# CLINICAL AND COST-EFFECTIVENESS OF A PERSONALISED GUIDED CONSULTATION VERSUS USUAL PHYSIOTHERAPY CARE IN PEOPLE PRESENTING WITH SHOULDER PAIN: A PROTOCOL FOR THE PANDA-S CLUSTER RANDOMISED CONTROLLED TRIAL AND PROCESS EVALUATION

**DOI:** 10.1101/2025.02.12.25322124

**Authors:** Sarah A Harrisson, Helen Myers, Gwenllian Wynne Jones, Ram Bajpai, Claire Bratt, Claire Burton, Rosie Harrison, Sue Jowett, Sarah A Lawton, Benjamin Saunders, David J Beard, Milica Bucknall, Rachel Chester, Carl Heneghan, Lucy Huckfield, Martyn Lewis, Christian D Mallen, Tamar Pincus, Jonathan L Rees, Edward Roddy, Danielle A Van der Windt

**Affiliations:** Primary Care Centre Versus Arthritis, School of Medicine, Keele University, Keele, Staffordshire, ST5 5BG, UK; Keele Clinical Trials Unit, School of Medicine, Keele University, Keele, Staffordshire, ST5 5BG, UK; Health Economics Unit, Department of Applied Health Sciences, University of Birmingham, Edgbaston, Birmingham, B15 2TT, UK; Nuffield Department of Orthopaedics, Rheumatology and Musculoskeletal Sciences (NDORMS), University of Oxford, OX3 7LD, UK; NHMRC Clinical Trials Centre, Faculty of Medicine and Health, The University of Sydney, Australia; School of Health Sciences, Faculty of Medicine and Health, University of East Anglia, Norwich Research Park, Norwich, NR4 7TJ, UK; Nuffield Department of Primary Care Health Sciences, University of Oxford, OX2 6GG, UK; Midlands Partnership University NHS Foundation Trust, Haywood Community Hospital, High Lane, Burslem, Stoke on Trent, Staffordshire, ST6 7AG, UK; School of Psychology, Highfield Campus, University of Southampton, Hampshire, SO17 1BJ, UK; NIHR Oxford Biomedical Research Centre, John Radcliffe Hospital, Oxford, OX3 9DU, UK; Haywood Academic Rheumatology Group, Midlands Partnership University NHS Foundation Trust, High Lane, Burslem, Stoke-on-Trent, Staffordshire, ST6 7AG, UK

**Keywords:** Musculoskeletal disorders, primary care, shoulder, physiotherapy, personalised care

## Abstract

**Introduction:** Musculoskeletal shoulder pain is a common reason for people to be treated in physiotherapy services, but diagnosis can be difficult and often does not guide treatment or predict outcome. People with shoulder pain cite a need for clear information, and timely, tailored consultations for their pain. This trial will evaluate the introduction of a personalised guided consultation to help physiotherapists manage care for individuals with shoulder pain.

**Methods and analysis:** This is a cluster randomised controlled trial to evaluate the clinical and cost-effectiveness of introducing a personalised guided consultation compared to usual UK NHS physiotherapy care. Physiotherapy services (n=16) will be randomised in a 1:1 ratio to either intervention (physiotherapy training package and personalised guided consultation incorporating a new prognostic tool) or control (usual care). 832 participants (416 in each arm) identified from participating physiotherapy service waiting lists aged 18 years or over with shoulder pain will be enrolled. Follow-up will occur at 3-time points: 6 weeks, 6 months and 12 months. The primary outcome will be the Shoulder Pain and Disability Index (SPADI) score over 12 months. Secondary outcomes include global perceived change of the shoulder condition, sleep, work absence and the impact of shoulder pain on work performance, healthcare utilisation and health-related quality of life (using the EQ-5D-5L). A multi-method process evaluation will investigate views and experiences of participants and physiotherapists, assess uptake, facilitators and barriers to delivery, and changes in factors assumed to explain intervention outcomes. Primary analysis of effectiveness will be by intention-to-treat, and a health economic evaluation will assess cost-utility of introducing the personalised consultation.

**Ethics and dissemination:** The trial received ethics approval from the Yorkshire & The Humber (South Yorkshire) Research Ethics Committee (REC reference: 23/YH/0070). Findings will be shared through journal publications, media outlets, and conference presentations. Supported by patient contributors and clinical advisors, we will communicate findings through a designated website, networks, newsletters, leaflets, and in the participating physiotherapy services.

**Trial registration:** ISRCTN: 45377604

**STRENGTHS AND LIMITATIONS OF THIS STUDY:**

- This large cluster randomised controlled trial will evaluate the effects and costs of a new intervention, comprising a scalable physiotherapy training package and a personalised guided consultation that incorporates a new prognostic tool, to improve care for patients with shoulder pain in physiotherapy services.
- A multi-method process evaluation, informed by predefined logic model, will assess the uptake, acceptability and delivery the intervention. This approach will allow for a comprehensive understanding of the perspectives and experiences of both participants and physiotherapists.
- The first phase will be an internal pilot designed to strengthen the trial by assessing recruitment, retention, and intervention uptake, increasing the likelihood of successfully delivering the full trial.
- A potential limitation of the cluster trial design is the increased risk of participation bias related to differential recruitment between trial arms.

## INTRODUCTION

Musculoskeletal shoulder pain is common, with 2− 3% of adults in the UK consulting with their GP for shoulder pain annually.^1,2^ Painful shoulder conditions affect sleep, social activities, work productivity, and result in increased healthcare use. Prognosis is highly variable, with approximately 40% of patients still experiencing pain six months after seeking treatment.^3–6^ Clinicians often face uncertainty when diagnosing the cause of shoulder pain,^7,8^ and physical examination results or imaging findings associated with assumed cause of shoulder pain do not always inform prediction of outcomes (prognosis) or response to treatment.^4,9^

Recent research emphasises the importance of factors beyond pathoanatomical diagnosis, such as sociodemographic variables, pain characteristics, general health, and psychological factors, in determining patient future outcomes.^10–12^ These prognostic factors assessed during consultations may help predict the course of shoulder pain and guide personalised treatment and self -management. Qualitative research in people with shoulder pain, including our own from an earlier phase of this programme of research,^8^ highlights the significant impact of the condition on daily life, work, and mood, and anxiety arising from uncertainty about prognosis, delays in diagnosis and unclear treatment options.^8,13,14^ There is a desire for clearer information, and timely, tailored consultations for patients with shoulder pain.^8^

In musculoskeletal pain research more broadly, there has been a move towards tailoring treatment options based on prognostic information, with people at high risk of persistent pain and disability being offered targeted or more extensive treatment, while offering advice and reassurance to those at low risk (stratified care).^15,16^ However, the results of trials comparing stratified care approach es have reported contrasting results with some demonstrating effectiveness or efficiency, while others find no benefit over usual primary care.^17,18^ To improve the design and evaluation of stratified or personalised interventions - especially those based on prognostic information – recommendations include refinement of risk prediction tools (e.g. by incorporating a wider range of prognostic factors), optimising intervention uptake, and better understand the challenges faced by clinical providers and recipients in delivering the intervention.^17^ Particularly in primary and community care, there has also been a call for approaches emphasising “de-medicalisation,” supporting people with shoulder pain to manage their own symptoms, and not just focus on the painful joint, but more holistically on the person living with a painful condition .^19^

The Prognostic And Diagnostic Assessment of Shoulder Pain (PANDA-S) trial is the final phase of a programme of research aiming to improve the management of shoulder pain in primary care (https://www.keele.ac.uk/panda-s/). A qualitative interview study, conducted as part of the PANDA-S research programme revealed disparities between people with shoulder pain and physiotherapists’ views towards shoulder pain consultations, indicating a need for improved patient-physiotherapist communication.^8^ Perspectives and experiences varied, including: (a) concerns from individuals with shoulder pain about the severity and impact of their condition, with limited opportunity to discuss these during consultations; (b) uncertainty or lack of confidence about diagnosis; (c) prognosis as a key concern, but with discrepancies between people with shoulder pain and physiotherapists regarding prognostic information offered; and (d) a perceived lack of information on treatment options, along with differing views regarding shared decision-making. Using a series of workshops with people with lived experience of shoulder pain and clinical advisors we co-designed a guided, personalised consultation and linked logic model (see Figure 1). This aimed to address key challenges, based on the principles of shared decision-making, effective reassurance, offering personalised advice about prognosis and treatment options, and building confidence to manage shoulder pain. The guided personalised consultation includes 3 components: (i) a pre-consultation leaflet for people with shoulder pain that they can use to prepare for a physiotherapy consultation and highlight their concerns and priorities; (ii) a prognostic tool developed as part of the PANDA-S programme to estimate likely future levels of pain and disability and inform the discussion about prognosis, and (iii) a consultation summary to be jointly completed by the person with shoulder pain and physiotherapist, summarising key decisions and signposting the individual to appropriate resources relevant to their condition and context.

**Figure 1.**
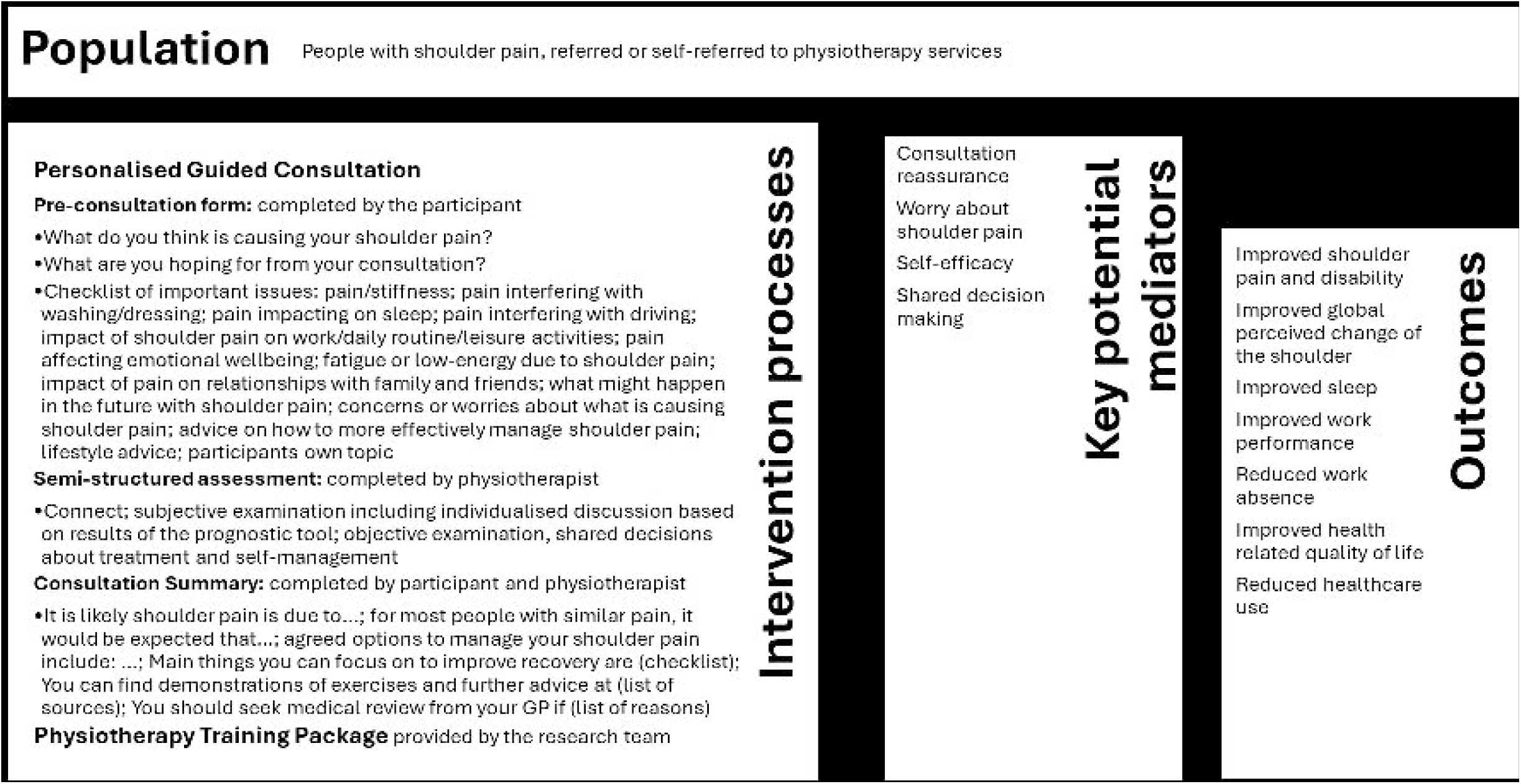
PANDA-S trial logic model

The PANDA-S trial aims to design and evaluate this personalised guided consultation for managing shoulder pain in physiotherapy services. This paper describes the protocol for the PANDA-S cluster randomised controlled trial (RCT) to test the clinical and cost-effectiveness of the introduction of the personalised guided consultation in physiotherapy services to support physiotherapists and people consulting for management of shoulder pain. The Standard Protocol Items: Recommendations for Interventional Trials (SPIRIT) checklist^20^ was used when writing this report.

## OBJECTIVES

The principal aim of the PANDA-S trial is to investigate the effectiveness of introducing the personalised guided consultation for people referred or self-referred to a physiotherapy service with shoulder pain on pain and disability over 12 months, compared to usual physiotherapy care.

The secondary aims are to:

- Investigate the effectiveness of the personalised guided consultation for people referred or self-referred to a physiotherapy service with shoulder pain and disability on perceived change of the shoulder condition, sleep, work absence and the impact on work performance, healthcare utilisation and health -related quality of life.
- Investigate the cost-effectiveness and cost-utility of introducing the personalised guided consultation compared to usual physiotherapy care.
- Investigate whether the effects of the personalised guided consultation are mediated by changes in self-efficacy, reassurance, worry, and shared decision-making.
- To evaluate the experiences of those participating with shoulder pain and physiotherapists regarding the delivery of the personalised guided consultation

## METHODS

### Design

This trial is designed as a cluster RCT with an internal pilot, process evaluation and economic evaluation in UK National Health Service (NHS) physiotherapy services to test the superiority of a personalised guided consultation (intervention arm) compared with care as usual (control arm).

### Settings and clusters

This cluster trial will take place in NHS physiotherapy services. For the purpose of the trial, and to minimise cross-contamination, a physiotherapy service will be defined as follows: A team of physiotherapists, working together at one clinic site for the large majority of their clinical practice. If staff work across multiple locations within a service, those locations will be considered as one cluster. People presenting with shoulder pain will be invited to participate in questionnaire data collection and interview studies. They will either receive the intervention or usual care or depending on the randomisation of the physiotherapy service they have been referred to.

A summary of the participant identification, invitation, and recruitment procedures for the study are outlined in the trial flow chart (see Figure 2).

**Figure 2.**
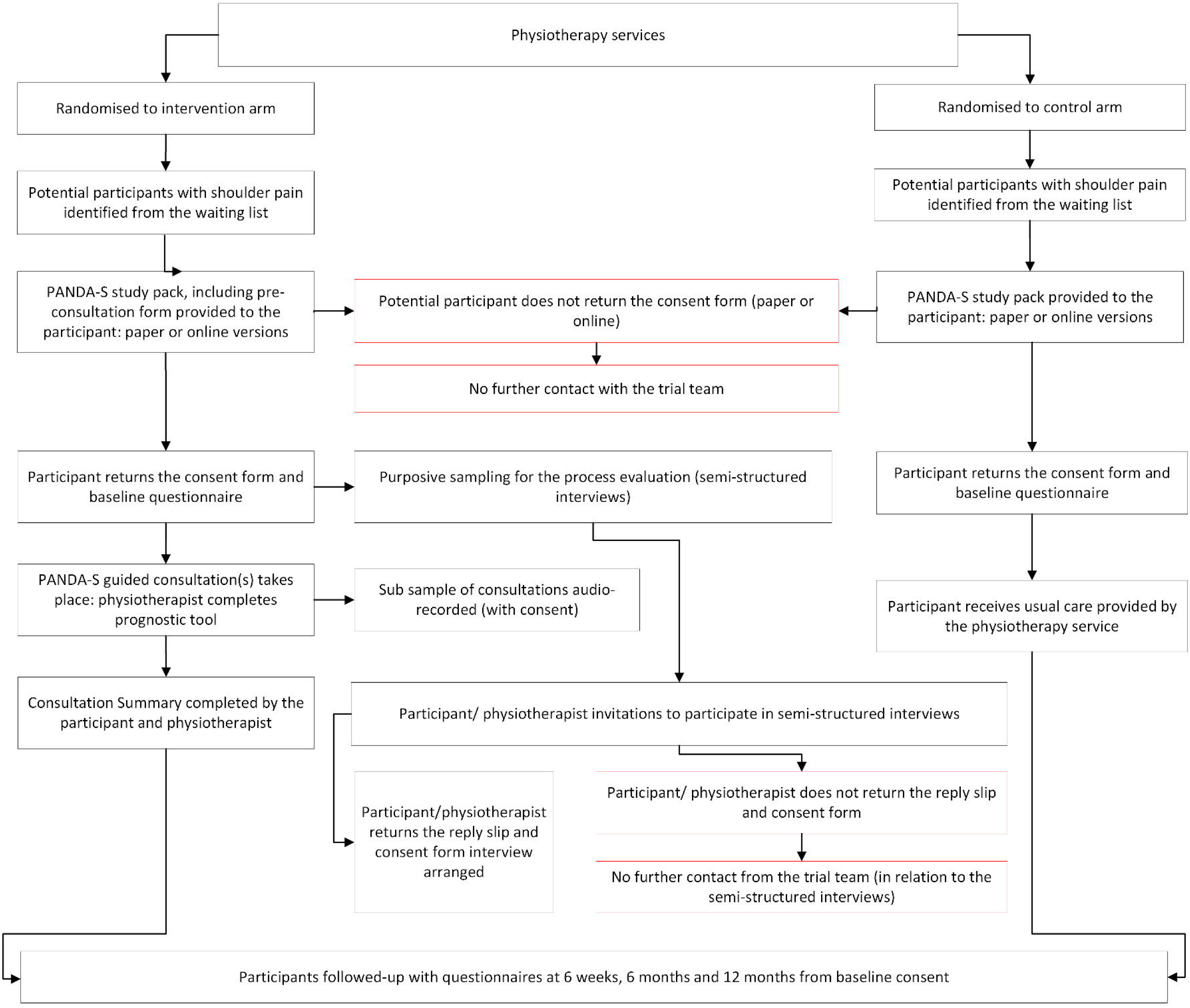
PANDA-S trial flow chart

### Inclusion criteria

People referred or self-referred to participating physiotherapy services, aged 18 years or over presenting with shoulder pain will be invited to participate.

### Exclusion criteria

People with shoulder pain will be excluded if they present to the physiotherapy service with symptoms or signs indicative of serious pathology (e.g. fractures, infection, inflammation, malignancy or referred pain from other sites (e.g. cardiac, hepatobiliary); have been referred for rehabilitation after shoulder surgery, have shoulder pain caused by stroke-related subluxation; have a diagnosis of inflammatory arthritis, including rheumatoid arthritis and polymyalgia rheumatica; have shoulder pain caused by cervical pathology or predominantly neck pain, or are considered by the staff triaging to be vulnerable (e.g. severe physical and/or mental health problems, dementia).

### Recruitment

We aim to recruit 832 people with shoulder pain from 16 services across England (see sample size calculation). Services will be identified through prior involvement in the wider research programme, response to an NIHR Research Delivery Network call for interest, or direct contact with Keele University Clinical Trials Unit (CTU) after learning about the trial via NIHR’s Open Data Platform.

After a service is identified, feasibility to take part will be assessed by discussion. This will consider: 1) referral numbers – the estimated number of eligible participants each month, 2) waitlist length – services with waitlists shorter than 4 weeks will be considered unsuitable, 3) service capacity – the ability to deliver the trial within the set recruitment period.

Participant recruitment started in November 2023 and is expected to close in March 2025. Potential participants will be identified from physiotherapy service waiting lists. Recruitment will be led by the physiotherapy service and integrated with their usual processes, using the PANDA-S eligibility checklist to support participant identification.

Potential participants will receive a study pack by post or an online link (via text message (SMS) or email). The packs will contain an invitation to participate, participant information sheet (PIS), a baseline questionnaire with consent form, pre-consultation form (for intervention arm only), and a reply-paid envelope (not included in the online invitation). Study packs will be sent directly from each physiotherapy service, with no reminders issued. Those who provide consent and their contact details will be considered enrolled. Completed packs will be returned to Keele CTU for processing.

### Randomisation

In line with a cluster RCT design, the NHS physiotherapy services will be the unit of randomisation. As the number of clusters will be small (envisaged to be up to 8 clusters per arm), we will use block randomisation stratified by the size of the physiotherapy service, based on the average number of patients treated for shoulder pain (large sites ≥150 per month, small sites < 150 per month). We will use block sizes of 2 within each stratum to randomly allocate the physiotherapy services to the intervention or control arms. The randomisation sequence will be computer generated using *blockrand* in R.^21^

To minimise risk of participation bias, all participants will receive the same PIS which informs participants that the study is looking at how the discussion between patients and physiotherapists can best be supported. The PIS does not refer to the trial having two arms (intervention and control). Participants will be asked to consent to participating in a study investigating delivery of care for shoulder pain by physiotherapists, consisting of the completion of self-report questionnaires (over a period of 12 months). Participants will be informed that they can withdraw from the study at any time without giving a reason , and that a decision to withdraw will not affect their current or future healthcare.

### Data collection

Trial processes will be managed by an online database: Research Electronic Data Capture (REDCap).^22^ Data collection will be carried out by self-completed questionnaires. Participants who complete a baseline questionnaire and consent to the study will be sent follow-up questionnaires at 6 weeks, 6 months, and 12 months. Questionnaires will either be paper-based or online depending on participant preference. Whilst the baseline study packs are sent directly from sites, follow-up questionnaires will be sent by Keele CTU. For paper-based questionnaires, reminders will be sent after 2 and 4 weeks. At each follow-up point, participants who do not respond to reminders and have provided a telephone number will be contacted for minimum data collection (MDC) by phone after 6 weeks with a MDC questionnaire after 8-weeks. Participants engaging online will receive reminders at weeks 1-4 inclusive. Vouchers will be sent with all follow-up questionnaires as a token of appreciation. Electronic Case Report Forms (CRFs) embedded in REDCap will be used to collect data from intervention sites. A schedule of the data to be collected is summarised in table 1.

**Table 1.** Data collection schedule in the PANDA-S trial.

| *Item* | *Description* | *B*<br>*L* | *6W* | *6M* | *12M* | *MDC* |
| --- | --- | --- | --- | --- | --- | --- |
| **Primary outcome measure** |  |  |  |  |  |  |
| *Severity of pain and disability* | SPADI total score (5 pain, 8 disability items, 0-10 NRS, 0-100) | ☐ | ☐ | ☐ | ☐ | ☐ |
| **Secondary outcome measures** |  |  |  |  |  |  |
| *Global perceived change* | Global Assessment of Change – one question, 5 options | ☐ | ☐ | ☐ | ☐ | ☐ |
| *Sleep* | Jenkins sleep questionnaire (4 items, 3 options) | ☐ | ☐ | ☐ | ☐ |  |
| *Work absence* | How many days off work have you had in the past 6 months (days/weeks)? | ☐ | ☐ | ☐ | ☐ | ☐ |
| *Work performance* | Two single item questions (0-10 NRS) | ☐ | ☐ | ☐ | ☐ | ☐ |
| *Healthcare utilisation* | Self-reported standardised items | ☐ | ☐ | ☐ | ☐ | ☐ |
| *Health-related quality of life* | EQ-5D-5L (5-items, 5 options each) | ☐ | ☐ | ☐ | ☐ | ☐ |
| **Anticipated mediators** |  |  |  |  |  |  |
| *Reassurance* | Consultation-based reassurance questionnaire (12 items, 7 options each) | ☐ | ☐ | ☐ | ☐ | ☐ |
| *Worry about shoulder pain* | Single item question (0 to 10 NRS) | ☐ | ☐ | ☐ | ☐ | ☐ |
| *Self-efficacy* | PROMIS (8 items, 5 options each) | ☐ | ☐ | ☐ | ☐ | ☐ |
| *Shared decision making* | CollaboRATE (3 items, 4 options) | ☐ | ☐ | ☐ | ☐ |  |
| **Characteristics of the shoulder pain condition** |  |  |  |  |  |  |
| *Shoulder pain related disability* | 0-10 NRS | ☐ | ☐ | ☐ | ☐ | ☐ |
| *Duration of shoulder pain episode* | Months | ☐ | ☐ | ☐ | ☐ | ☐ |
| *Acute or traumatic onset* | Single item question (yes /no) | ☐ | ☐ | ☐ | ☐ | ☐ |
| *History of shoulder pain* | Single item question (3 options - no, yes once, yes multiple times) | ☐ | ☐ | ☐ | ☐ | ☐ |
| **Psychological and behavioural factors related to shoulder pain** |  |  |  |  |  |  |
| *Avoidance of activities* | Single item question (4 options) | ☐ | ☐ | ☐ | ☐ | ☐ |
| *Belief that movement is harmful* | Single item question (0-10 NRS) | ☐ | ☐ | ☐ | ☐ | ☐ |
| *Confidence in managing shoulder pain* | Single item question (0-10 NRS) | ☐ | ☐ | ☐ | ☐ | ☐ |
| *Treatment expectations* | Single item question (0-10 NRS) | ☐ | ☐ | ☐ | ☐ | ☐ |
| **Lifestyle and general health** |  |  |  |  |  |  |
| *Pain elsewhere* | Two single item questions (yes /no) | ☐ | ☐ | ☐ | ☐ | ☐ |
| *Presence of diabetes mellitus* | Single item question (yes /no) | ☐ | ☐ | ☐ | ☐ | ☐ |
| *Physical activity* | Single item question (7 options) | ☐ | ☐ | ☐ | ☐ | ☐ |
| *Symptoms of anxiety* | GAD-2 (2 items, 4 options) | ☐ | ☐ | ☐ | ☐ | ☐ |
| *Symptoms of depression* | PHQ-2 (2 items, 4 options) | ☐ | ☐ | ☐ | ☐ | ☐ |
| **Sociodemographics** |  |  |  |  |  |  |
| *Age* | Date of birth | ☐ | ☐ | ☐ | ☐ | ☐ |
| *Sex* | Male / Female / Prefer not to say | ☐ | ☐ | ☐ | ☐ | ☐ |
| *Education* | Qualifications | ☐ | ☐ | ☐ | ☐ | ☐ |
| *Ethnicity* | Single item | ☐ | ☐ | ☐ | ☐ | ☐ |
| *Current work situation* | Single item | ☐ | ☐ | ☐ | ☐ | ☐ |
| *Job title* | Open question | ☐ | ☐ | ☐ | ☐ | ☐ |
| *Area level of deprivation* | Derived from postcode | ☐ | ☐ | ☐ | ☐ | ☐ |
| *Health Literacy* | Single item literacy screener (5 options) | ☐ | ☐ | ☐ | ☐ | ☐ |
| *Body Mass Index* | Self-reported height and weight | ☐ | ☐ | ☐ | ☐ | ☐ |

Abbreviations: 6W= 6 weeks, 6M= 6 months, 12M= 12 months, BL= Baseline, MDC = Minimum Data Collection. GAD-2, Generalised Anxiety Disorder questionnaire – 2 items. PHQ-2, Patient Health Questionnaire - 2 items. PROMIS, Patient-Reported Outcomes Measurement Information System. SPADI, Shoulder Pain and Disability Questionnaire.

The primary outcome is the Shoulder Pain and Disability Questionnaire (SPADI)^23^ score over 12-month follow-up, where a higher total score (0-100) indicates worse pain and/or disability.

Secondary outcome measures will assess: 1) overall patient experience (using global perceived change in shoulder pain, 2) sleep using the Jenkins Sleep Questionnaire,^24^ 3) work absence and the impact of shoulder pain on work performance using the single item work performance scale,^25^ 4) healthcare utilisation and 5) health-related quality of life using the EQ-5D-5L.^26,27^

Measures of anticipated key mediators, informed by the intervention logic model, will assess: 1) reassurance using the Consultation-based Reassurance Questionnaire,^28^ 2) worry about shoulder pain in the past week, 3) self-efficacy (8-item Patient-Reported Outcomes Measurement Information System (PROMIS) scale for Self-Efficacy (managing symptoms of chronic conditions),^29^ 4) participants’ experience of shared decision making using the CollaboRATE measure.^30^

The baseline questionnaire will be used to measure the following prognostic factors: Shoulder pain characteristics (shoulder pain intensity,^31^ shoulder pain related disability, duration of shoulder pain episode, acute or traumatic onset, history of shoulder pain). Psychological and behavioural characteristics (avoidance of activities, belief that movement is harmful, confidence in managing shoulder pain, treatment expectations).^31^ General health and lifestyle (presence of pain elsewhere,^32^ diabetes mellitus, level of physical activity^31^). Anxiety and depression using the Generalised Anxiety Disorder – 2 item (GAD-2)^33^ questionnaire for anxiety and the Patient Health Questionnaire – 2 items (PHQ-2) for depression.^34^ These factors were chosen based on previous research, input from clinical advisors, and insights from people with lived experience of shoulder pain.

Age, sex, body mass index (BMI) ethnicity, education (to identify highest qualification), health literacy,^35^ current work situation, most recent paid job title, and psychosocial work environment will be measured to enable a description of the population.

### Trial intervention

We combined best practice guidance^36^ with the results from a qualitative interview study ^8^ and a series of workshops involving patient contributors and clinical advisors to co-design and develop the intervention (the personalised guided consultation and associated training package for Physiotherapists) informed by the Medical Research Council (MRC) framework for complex interventions.^37^ The guided consultation is based on the principles of shared decision-making,^38^ to include effective reassurance, building confidence (self-efficacy) and provision of personalised advice and treatment, and comprises of the following 3 elements:

1. **Pre-Consultation Form** will be sent to participants in advance of the consultation (with the baseline questionnaire) on which they can highlight their concerns and priorities about their shoulder pain and expectations for the consultation.
2. **A Semi-Structured Assessment** to guide physiotherapists in covering key elements of the consultation:

- Subjective examination, which will include an assessment of items for the prognostic tool; an assessment of ideas, concerns and expectations (ICE)^39,40^ as highlighted by participants on the pre-consultation form; an assessment of personal values in terms of their shoulder pain and disability
- Standardised physical examination , as per usual care
- Personalised discussion of expected course of shoulder pain and disability, based on individualised results of the prognostic tool and personal values
- Discussion of treatment and self-management options, tailored to the identified concerns, priorities, expectations, values, results of physical examination, and individual prognosis. The prognostic tool included the items with prognostic value collected in the baseline questionnaire and allows an individualised prediction of the level of pain and disability (SPADI score) at 3 and 6 months displayed as a bar chart.
3. **A Consultation Summary** will be jointly completed by participant and physiotherapist to support shared decision making. The summary will outline shared decisions regarding treatment and self-management options, for the patient to keep for reference following the consultation.

Physiotherapists will be provided with training in how to deliver the intervention using the pre-consultation form, assessment and consultation summary. All participating physiotherapists will complete a 4-hour training programme, which will be delivered either online or face-to-face, depending on the preference of the physiotherapy service. The training will provide content on the rationale for the guided consultation, core components of the consultation and key skills to support participating physiotherapists to work towards making shared decisions. A comprehensive package of online resources will be made available to participating physiotherapists working in services randomised to the intervention arm.

For services randomised to the control arm, which will continue to offer care as usual, training will involve a short online session to explain the objectives and overall design of the trial, the procedures for eligibility screening and for sending out study packs to potentially eligible participants (this same session will also be offered to triage teams in services randomised to the intervention arm).

### Process evaluation

The process evaluation will explore and assess factors influencing the delivery of the intervention and provide important context for interpreting the trial results. It will examine the credibility and acceptability of the intervention, gathering patient participants’ views on their satisfaction with the care they received, the perceived impact on outcomes, and their experiences of taking part in the trial. It will also capture physiotherapists’ perspectives on delivering the guided consultation, including their views on the intervention’s effectiveness and its impact on their approach to shoulder pain management. These qualitative data will help identify key factors shaping the intervention’s delivery and outcomes. A multi-method approach will be used which will include semi-structured interviews with participants and physiotherapists, audio-recording of the guided consultation, analysis of data collected during the consultation using CRFs, and participant questionnaires.

### Semi-structured interviews

Semi-structured interviews focusing on how the intervention is used and understood and guided by the elements and principles underpinning the intervention will be carried out with patient participants and physiotherapists:

- Patient participants (up to n=20) will be selected to represent different prognostic profiles, ages, genders, work statuses, and geographic areas. A sample of these interviews will be conducted in the first 6 months of recruitment to help interpret results from the internal pilot phase and refine the intervention or trial processes, such as refinements to physiotherapist training. Interview topics will cover experience of recruitment, questionnaire completion, views on discussions about shoulder pain diagnosis, prognosis, and management of shoulder pain during consultations, perceptions of advice given, how decisions were made, reassurance provided, and overall confidence to manage shoulder pain and satisfaction with their treatment.
- Interviews with physiotherapists (approx. n=10) will focus on their views regarding the content and usefulness of training materials, changes in confidence in managing shoulder pain, assessing and discussing prognosis, and providing reassurance. Their views on perceived barriers and facilitators to delivering the guided consultation will also be explored.

Patient participants who agree to be contacted for an interview in their baseline questionnaire will be sampled to ensure variation in characteristics such as reported pain severity, ethnicity, age, gender, work status, and geographic location (across different physiotherapy services). Patients will receive an invitation letter and information leaflet detailing the interview purpose and information about confidentiality, data storage, and archiving. A reply slip will also be included for them to return to indicate interest and provide contact details. A research team member will then contact the participant to schedule the interview (e.g., either by phone, or videoconference). Before the interview, verbal consent will be audio-recorded with consent reaffirmed at the end.

Physiotherapists will be invited to participate in interviews as part of their involvement in the trial, with sampling based on gender, work location, and years of clinical experience. They will receive an interview invitation and information leaflet by email. A research team member will contact them to arrange the interview, which will take place using either phone or video call. Consent will be obtained in the same manner as for patients.

The final number of interviews will be guided by inductive thematic saturation, where data collection stops once no new themes or codes are identified.^41^ The semi-structured interviews will allow the interviewer, trained in qualitative methods, to explore additional relevant topics as they arise. The topic guides will be revised iteratively during data collection and analysis based on ongoing findings.

### Audio-recording of consultations

Up to 10 guided consultations will be recorded to assess the use of the 3 components of the intervention, specifically whether it is delivered according to protocol, including effective reassurance, personalised self-management advice, and treatment based on the individual needs of the patient and their estimated prognosis.

Patient participants who agree to be contacted for consultation recordings in their baseline questionnaire will be selected using the same criteria as for the semi-structured interviews. These participants will receive an invitation letter, information leaflet, and consent form. For those participants who consent to their consultation being recorded, the research team will liaise with the physiotherapy service to confirm the date of the consultation. The physiotherapist, who will also complete a consent form, will be informed. Both participant and physiotherapist consent will be obtained before the consultation is recorded.

The physiotherapist will reaffirm the patient’s consent and use a secure, password-protected digital recorder to record the consultation. Afterward, the recording will be couriered to Keele CTU for upload to a secure SharePoint location, and the original file will be deleted. If consent is not provided or is withdrawn, or a suitable room is unavailable, the consultation will not be recorded.

### Data collected during the personalised guided consultation

Descriptive analysis of the CRFs, Consultation Summary, and patient questionnaires will provide quantitative data on the following: uptake of the intervention by physiotherapists, consultation duration, and the extent to which treatment and self -management options align with patients’ expressed needs, prognostic factors, and predicted future levels of shoulder pain and disability. The patient questionnaires (at baseline, 6 weeks, and 6 months) will also capture data on potential mediators (reassurance, worry, self-efficacy, and shared decision - making), helping to explore how these factors may influence the intervention’s impact on patient outcomes.

### Internal pilot

The internal pilot will aim to assess recruitment, retention and intervention uptake, test trial procedures and explore intervention acceptability. It will form the first phase of the main trial recruiting at least 200 participants from 4 to 6 physiotherapy services over 6 months. Recruitment will not be stopped at the end of the internal pilot period but there will be the opportunity while recruitment is ongoing to potentially revise trial procedures for the main trial. To improve intervention uptake, actions may include: identifying barriers to adoption with physiotherapists, revising training materials and offering refresher courses, providing mentorship and supervision from senior team members. A priori defined success criteria will be used to assess progress during the internal pilot phase (see Table 2). These criteria are similar to stop/go criteria, but their purpose is to inform discussions with the Programme Steering Committee (PSC – see Trial monitoring).

**Table 2.** A priori defined criteria to assess progress during the internal pilot phase of the PANDA-S trial.

|  | *Continue as planned* | *Implement remedies & continue, with protocol amendments as needed* | *Discuss the feasibility of continuing with the Programme Steering Committee* |
| --- | --- | --- | --- |
| *Recruitment* | ≥ 80% of the target participants by 6 months (n ≥ 160) | ≥ 50% <80% of the target participants by 6 months | <50% of the target participants by 6 months |
| *Follow-up rate (6 weeks)* | ≥ 70% retention | ≥ 50% <70% retention | <50% retention |
| *Risk of participation bias (baseline similarity)* | <15% difference in participant characteristics | >15% difference in participant characteristics | - |

### Analysis

Data will be reported according to the reporting guidelines for randomised clinical trials: CONSORT (2010) statement^42,43^ including extensions to cluster randomised trials^44^ and pragmatic trials.^45^ A CONSORT style flow diagram will be used to show the flow of participants through the trial, including reasons (where given) for withdrawal at both the physiotherapy service and individual participant level. These will be recorded on REDCap. Analysis will be conducted by a statistician who will remain blind to allocation of physiotherapy clusters to either intervention or control arm.

### Internal pilot analysis

Data from the internal pilot phase will be analysed using descriptive methods.

### Sample size calculation

The sample size for the trial is based on previous studies using the SPADI^18,46^ and using data from the PANDA-S cohort study.^32^ The trial aims for 90% power to test the superiority of the intervention to usual care by physiotherapists for patients with shoulder pain. The calculations assume a 5% type I error (two-tailed) and aim to detect an effect size of 0.30 (8-10 point difference, SD 24-30) in the primary outcome.^47^ The calculation takes into account the clustering of individual participants by service (ICC 0.02)^48^ and likely loss to follow-up of 25% over the 12-month follow up (inflationary effect on the sample size), repeated measurements^49^ and adjustment for baseline SPADI scores (deflationary effect). To achieve this, 416 participants per arm, or 832 in total, are required.

### Main trial analysis

Baseline characteristics of the physiotherapy service clusters, and individual participants will be described for each trial arm. This includes the primary outcome, secondary outcomes, process measures, predicted SPADI scores, and the proportion of participants predicted to score 20 or lower on the SPADI, indicating recovery.

The primary intention-to-treat (ITT) analysis will use a hierarchical linear mixed regression model to compare offering the intervention with usual care on SPADI total scores over the 12 months follow-up. This model will account for repeated measures within individuals and clustering within physiotherapy services, using available data from all-time points and assuming missing data are random. Analyses will adjust for baseline SPADI score, age, sex, and physiotherapy cluster size. A non-responder analysis will assess the risk of attrition bias. Data for individuals who withdraw consent will be included up to the point of withdrawal.

We will report mean differences and standardised effect sizes (relative to the overall standard deviation (SD)) for the primary outcome. The number (percentage) of participants in each trial arm scoring 20 or lower on the SPADI at 6- and 12-months follow-up will also be reported, which will be used to calculate the Number-Needed-to-Treat (NNT).

The model parameters will primarily be estimated using maximum likelihood estimation (MLE). To address potential issues due to the small number of clusters, restricted maximum likelihood estimation (RMLE) will also be used as a sensitivity analysis. This will include evaluations with and without the Kenward-Roger approximation correction^50^ for standard error and degrees of freedom in estimating fixed effects.

Analysis of secondary outcomes will be carried out using an ITT approach , using a linear mixed model for numerical outcomes and generalised mixed logistic models for categorical outcomes. The models will be adjusted for baseline SPADI, age, and sex (participant-level), and physiotherapy service cluster size. A sensitivity analysis will be considered if there is baseline imbalance between arms, with additional adjustment for baseline characteristics that are >10% different for categorical variables, or >2 times the standard error for numerical variables. For work absence, performance and healthcare utilisation, the analysis will focus 6- and 12-month follow-up data, as these measures are not collected at the 6-week follow-up.

Sensitivity analyses will be carried out to provide an unbiased estimate of intervention effect for participants who received the guided consultation as per protocol. Exploratory subgroup analyses will be conducted for the primary outcome to explore the effectiveness of the intervention. These analyses will be carried out in participants with 1) long (> 6 months) versus short baseline pain duration; 2) having received previous treatment for their shoulder condition, or not; 3) low versus increased levels of distress (symptoms of anxiety and depression); and 4) those living in areas with low versus high deprivation .

### Health economic analysis

A within-trial economic evaluation will assess the cost-effectiveness and cost-utility of the intervention versus usual care for shoulder pain over 12 months. Incremental analysis will estimate the cost per additional point improvement on the SPADI (cost-effectiveness) and per quality-adjusted life year (QALY) gained (cost-utility) from both NHS and societal perspectives. The EQ-5D-5L (collected at baseline, 6 weeks, 6 and 12 months) will be used to generate QALYs. Healthcare resource use information will capture costs related to delivery of the guided consultation and training package, usual care, medicines use and any additional primary and secondary healthcare required for shoulder pain. Information on time off work will capture productivity losses.

Mean costs and outcomes will be compared between trial arms, with missing data handled using multiple imputation. Statistical analysis will be done on an ITT basis, adjusting for clustering and baseline variables. Incremental cost-effectiveness ratios will be calculated, and uncertainty will be examined by estimating 95% CIs and cost-effectiveness acceptability curves (CEACs), which link the probability of the intervention being cost-effective to a range of potential threshold values thatthe health system may be willing to pay per additional QALY gained. Analysis will adopt methods reflecting the cluster randomised nature of the trial, by using a regression -based model of net benefits, with physiotherapy service as the cluster identifier. Dependent variables in the multilevel models will include costs, QALYs and net monetary benefits (NMB) and model co-efficient estimates of differences in these variables will be used as part of the incremental analysis.

A decision-analytical model will estimate long-term cost-utility of the intervention, using data from the trial, clinical expertise within the team, the PANDA-S cohort study^31^ and estimates from the literature on shoulder pain prognosis, costs and quality of life. Sensitivity analyses will explore parameter uncertainty, with probabilistic sensitivity analysis to generate CEACs. Value of information analysis will assess the value of additional data to reduce uncertainty.

### Process evaluation analysis

Semi-structured interviews will be video or audio-recorded, transcribed, anonymised, and analysed using two stages. First, an inductive thematic analysis^51^ will identify themes, followed by mapping these themes onto two theoretical frameworks:

- *Normalisation Process Theory* (NPT),^52^ which explores how interventions are adopted and integrated into clinical practice through four components: coherence, cognitive participation, collective action, and reflexive monitoring.
- *COM-B model*^53^ which examines behaviour change of both those delivering (physiotherapists) and receiving the intervention (people with shoulder pain) through capability, opportunity, and motivation, extending the Theoretical Domains Framework (TDF).^54^

A coding framework will be developed from early transcripts, and then applied to the remaining interviews, and refined iteratively. Researchers will compare the views of participant and physiotherapists, analysing data for consistency and variation, and aligning themes with NPT and COM-B components.

Audio-recorded consultations will undergo in-depth analysis using theme-oriented discourse analysis.^55^ This method will focus on the linguistic features and interactional strategies used by patients and clinicians during consultations. Researchers will transcribe and analyse content, paralinguistic features, such as pauses and intonation, and discursive strategies employed by people with shoulder pain and physiotherapists, to understand how the intervention is used and how participants engage with it. The team will discuss and agree on interpretations across consultations.

Data from CRFs and consultation summaries will be analysed using descriptive methods and will provide information on the physiotherapists opinions on the shoulder pain presented by patients. The analysis will assess the extent to which the three components of the guided consultation has been taken up, the time spent, and the type of treatments and self-management resources offered. Physiotherapist comments on barriers and/or facilitators during consultations will be reviewed and categorised according to themes from the interviews and recordings.

### Mediation analysis

Data from patient questionnaires will be analysed as potential mediators of the guided consultation’s effect. Mediation models for longitudinal data, using structural equation modelling or the outcomes framework, will estimate how changes in mediators explain the intervention’s impact on the primary outcome through indirect pathways.

### Trial monitoring

The PSC was appointed and approved by the funders to oversee the scientific conduct of the programme grant. Providing independent oversight of the trial, the PSC includes a chair, patient partner, senior statistical representative, and two experienced clinical academics with relevant expertise. The committee met during the set-up of the trial and will meet every 6 months thereafter, for the duration of the trial. Given the low risk of the trial - the PSC and funders agreed that an independent Data Monitoring Committee (DMC) was not needed. Data monitoring responsibilities have therefore been adopted by the PSC.

Reporting to the PSC, a Trial Management Group (TMG) has been formed and comprises the chief investigator, associate investigators, co-applicants, trial managers from Keele University Clinical Trials Unit (CTU) and statistician plus other stakeholders as required. In addition to the management and monitoring of the trial, the TMG is responsible for optimal delivery of the trial, analysis and interpretation of the results.

### Safety reporting

In the PANDA-S trial, the guided consultation involves introduction of pre-consultation form, a semi-structured assessment, consultation summary documents, and personalised information and advice. Adverse events (AEs) are expected to be rare and minor. Events such as temporary pain or discomfort from physiotherapy assessments and hospital treatments for planned shoulder surgeries will not be recorded as highly unlikely to be related to the intervention. However, if a participant becomes distressed during the delivery of the guided consultation and the physiotherapist deems this related to the intervention, it will be reported as AE. In the event of either an AE or serious adverse events (SAEs) there are systems in place to inform the CTU. Any SAEs will be reported to the research ethics committee (REC) within the relevant time frame, and to the TSC as appropriate.

### Ethical considerations Ethics approval

This trial was provided with ethical approval by the Yorkshire & The Humber (South Yorkshire) Research Ethics Committee (REC reference: 23/YH/0070) on 28^th^ April 2023.

### Participant consent

Informed consent for physiotherapy services to participate will be provided by the physiotherapy service lead. A Model Agreement for Non-Commercial Research (mNCA)^56^ will be signed by the sponsor at Keele University prior to randomisation. Written informed consent for data collection will be obtained from eligible patients presenting with shoulder pain who are willing to participate in the trial.

### Regulatory compliance

The PANDA-S trial will follow Good Clinical Practice (GCP) principles and the UK Policy Framework for Health and Social Care Research. Keele University, as the sponsor, has a Health and Social Care Research (HSCR) Quality Management System (QMS) with Standard Operating Procedures (SOPs) in place which will be adhered to in the conduct of the trial. An independent external audit may be carried out by the sponsor. For quality assurance purposes, trials supported by the sponsor may be subject to an independent audit. The Head of Project Assurance at Keele University (the trial sponsor) can be contacted by.

### Modification of the protocol

The trial sponsor will be notified of all amendments to the protocol.

### Protocol compliance

Deviations from the protocol and GCP will be documented. Keele CTU will implement corrective and preventative actions where appropriate. The Chief Investigator will take responsibility for these actions, with approval from the PSC if necessary.

### Data protection and patient confidentiality

All information collected during the trial will be kept strictly confidential and securely managed by Keele University through the CTU. Keele CTU adheres to General Data Protection Regulation ^57^ regulations and maintains a duty of confidentiality. All sensitive and personal electronic data will be stored in the CTU’s secure virtual network, requiring two-factor authentication for access. User roles and permissions will limit access to specific data and operations. After data collection has been completed, all data will be anonymised and cannot be linked to identifiable participants. Hard copies, such as consent forms, will be securely stored in lockable filing cabinets at Keele CTU.

### Post trial care

All participants will receive care from their treating physiotherapist. The trial will not provide treatment or make recommendations regarding specific diagnostic procedures or clinical treatments, either during or after its completion.

### Access to the final trial dataset

Keele University, a member of the UK Reproducibility Network, is committed to the principles of the UK Concordat on Open Research Data. Keele CTU has a longstanding commitment to sharing trial data to enhance research reproducibility and maximise benefits for patients, the public, and the health and social care system. All data are available on following the School of Medicine at Keele University data request process by contacting the corresponding author and.

### Patient and public involvement

Having already provided input into the development of the PANDA-S intervention, patient contributors will advise on procedures integral to the success of the trial. Meetings with patient partners will be arranged at critical time points during the trial to discuss the following key areas:

- Recruitment and retention
- Acceptability, credibility and uptake of the intervention
- Dissemination of findings

### Dissemination

Trial results will be presented at local, national, and international conferences and published in free-access peer-reviewed journals. Following publication, further dissemination will occur through updates on Keele University’s website (https://www.keele.ac.uk/), summaries for participating physiotherapy services, and communication with related patient groups. To ensure these findings inform policy and practice, a dissemination plan has been developed based on National Institute for Health and Care Research (NIHR) “Push the Pace” guidance.^58^ To implement this plan, the PANDA-S team will seek advice from Keele University’s Impact Accelerator Unit, which has a designated team working towards accelerating the uptake of research into practice.

## DISCUSSION

The PANDA-S trial addresses key challenges in managing musculoskeletal shoulder pain within primary care services: difficulties in communication between patients and healthcare professionals regarding concerns about shoulder pain and expectations of management highlighted by qualitative research; the uncertainty in identifying and discussing individual prognosis; and agreeing advice and treatment options tailored to individual patients’ concerns, shoulder pain characteristics and prognosis. To tackle these challenges, the trial will evaluate the effectiveness of a personalised guided consultation that incorporates a co-designed prognostic tool. The trial’s main findings on the effectiveness and cost-effectiveness of introducing the personalised guided consultations for people with shoulder pain in physiotherapy could have significant implications for patients, the NHS, and policy. If the guided consultations—through factors such as shared decision -making, improved confidence in symptom management, and effective reassurance—lead to better outcomes compared to the control arm, this research will provide valuable insights. It will advance the field of shoulder pain and musculoskeletal care while contributing to the evidence base needed to reduce unwarranted variation in physiotherapy care.

## Supporting information

SPIRIT Checklist

## Data Availability

All data are available on following the School of Medicine at Keele University data request process by contacting the corresponding author and.

## Disclaimer

The views expressed in this publication are those of the authors and not necessarily those of the NHS, the NIHR or the Department of Health and Social Care.

## Author Statement

DvdW, GW-J, HM, MB, DB, CB, RC, CH, LC, LH, RH TP, BS conceptualised and developed the idea for the trial. MB, RB and ML provided statistical expertise in clinical trial design. DvdW, SH, HM, RB, BS, RH, SL provided time and expertise with data collection and project administration. SH, GW-J, DvdW, HM, BS, RB, RH, SJ wrote the first draft of the manuscript. All authors contributed to the refinement of the protocol, critically edited the manuscript, read and approved the final manuscript.

## Acknowledgements

The authors acknowledge the support of NIHR Research Delivery Network, specifically Joe Goldby and Natasha Upton at West Midlands, and Keele Clinical Trials Unit for their support in delivering and hosting this research ; the patient partners and clinical advisors, for their valuable contributions to the development of this trial; Dr Elaine Nicholls at Keele University for leading the process of randomising physiotherapy services; physiotherapy services and research delivery teams at our participating sites (Airedale NHS Foundation Trust, Birmingham Community Healthcare NHS Foundation Trust, Dorset Community NHS Trust, Hampshire and Isle of Wight Healthcare NHS Foundation Trust, Hereford and Worcester Health and Care NHS Foundation Trust, Hertfordshire Community NHS Trust, Horder Healthcare, Gloucestershire Hospitals NHS Foundation Trust, Great Western Hospitals NHS Foundation Trust, Leeds Community Healthcare NHS Trust, Lewisham and Greenwich NHS Trust, Medway Community Healthcare, Midlands Partnership University NHS Foundation Trust, Wirral Community Health and Care NHS Foundation Trust) for their essential contributions.

